# Whole genome sequencing-based analysis of genetic predisposition to adult glioblastoma

**DOI:** 10.1101/2025.01.16.25320661

**Authors:** Mark P. van Opijnen, Devin R. van Valkengoed, Joep de Ligt, Filip Y.F. de Vos, Marike L.D. Broekman, Edwin Cuppen, Roelof Koster

## Abstract

**Background:** Glioblastoma is most commonly reported in the second (pediatric form) and seventh (adult form) decade of life. Pathogenic germline variants (PGVs) and its association to late onset glioblastoma remains unclear. This study aimed to investigate the genetic predisposition to adult glioblastoma.

**Methods:** We performed an in-depth analysis of whole genome sequencing (WGS) data of tumor-normal tissue pairs of 98 glioma WHO grade 4 patients for potential presence of PGVs, in a comprehensive set of 170 genes associated with cancer predisposition. All candidate pathogenic events were also assessed for second-hit somatic events.

**Results:** In 11 patients (11%), PGVs were observed that were considered relevant by clinical experts in the context of glioblastoma. In these patients, 13 PGVs were found in genes known for a strong association with familial glioblastoma (*MSH6* (3x), *PMS2* (5x), *MSH2*, *TP53*, *NF1* and *BRCA1*) or with medulloblastoma (*SUFU*). In eight of these patients (73%) causality was supported by a second (somatic) event and/or a matching genome-wide mutational signature.

**Conclusions:** Germline predisposition does also play a role in the development of adult glioblastoma, with mismatch repair deficiency being the main mechanism. Our results do illustrate benefits of tumor-normal WGS for glioblastoma patients and their relatives beyond the identification of potentially actionable mutations for therapy guidance.

**Key points:**

1. Pathogenic germline variants occur in more than 10% of adult glioblastoma
2. Mismatch repair deficiency is the main predisposition mechanism
3. Pathogenic germline variants could be used for (targeted) treatment selection

**Importance of the Study:** The hereditary of adult glioblastoma is still largely unexplored. With the option of broad molecular testing, it is crucial that clinicians are aware of the a priori probability of finding germline predisposition in a glioblastoma patient. Here, we studied the genetic predisposition to adult glioblastoma in an unselected, average cohort. We observed that pathogenic germline variants occurred in about 1 out of 10 patients, with mismatch repair deficiency being the main predisposition mechanism. This information should be kept in mind when broad molecular testing, like WGS, is discussed with the patient. Clinicians and patients should discuss the probability of finding evidence of heredity of the tumor and potential consequences for relatives.

## Introduction

Glioblastoma, a primary brain tumor, is the most common and most aggressive malignant brain tumor in adults. Despite intensive treatment consisting of surgical resection followed by radiotherapy with concurrent and sequential chemotherapy, the prognosis remains poor with a median survival of 15 months.^1, 2^ One of the contributing factors challenging effective treatment strategies is the inter- and intratumoral heterogeneity of this devastating disease.^3^ This becomes also apparent in the complexity revealed by genomics^4^ and single-cell RNA sequencing.^5, 6^ Nevertheless, genomic analysis of the tumor is considered a promising technological development that could enable personalized treatment strategies. The most comprehensive approach for genomic analysis is whole genome sequencing (WGS), which has been clinically validated for diagnostic purposes.^7, 8^ WGS is not yet widely used in routine settings, especially for glioblastoma, mostly because of lack of evidence of clinical utility, costs, or both. Therefore, we have initiated the GLOW trial, a clinical study to explore potential added value of WGS for recurrent glioblastoma patients.^9^ As WGS analyses typically include a control normal DNA sample (e.g. from blood) to distinguish somatic variants (acquired in the tumor cell) from germline variants (inherited), they may also reveal potential genetic predisposition to glioblastoma. This knowledge might be relevant to patients and their relatives, and the presence of familial predisposition is often an important question in the consulting room. Furthermore, variants in several predisposition genes are increasingly important for (immune- or targeted) therapy selection.^10–16^

In contrast to other (sub)types of cancer, for instance breast cancer and colon cancer, but also to pediatric gliomas, the prevalence of heredity in adult glioblastoma patients is still largely unexplored, mainly due to lower incidence and limited datasets that are available to investigate this topic.^17–19^ In general, an estimate of approximately 5% of all glioma patients have a positive family history for glioma, with twofold to elevenfold increased incidence ratios in those families.^20–22^ These cases show similarity to sporadic cases in terms of demographics (age, gender), morphology and tumor grade, and penetrance of hereditary glioma is suggested to be low.^23^ Hereditary glioblastoma, also called familial glioblastoma, caused by single-gene hereditary disorders is very rare^24^ and often involves predisposition of a range of tumor types. Current knowledge is limited to a few syndromes including neurofibromatosis type 1 (*NF1* mutation, autosomal dominant), Li Fraumeni syndrome (*TP53* mutation, autosomal dominant), Turcot syndrome type 1 (mismatch repair genes [MLH1 & PMS2] mutations, autosomal dominant) and Lynch or constitutional mismatch repair deficiency (mismatch repair genes mutations, autosomal dominant [Lynch] or recessive [constitutional mismatch repair deficiency]).^25–27^ Furthermore, in enriched cohorts (i.e. selected for personal and/or family history) pathogenic variants in *BRCA 1* and *2*, *CHEK2*, *HERC2*, *MUTYH*, *NF1*, *POT1* and *TERF2* have been associated with glioblastoma^20, 28–30^, although their contribution to glioblastoma development remains unclear, since second-hit somatic variants were not observed for many.^29^ Apart from these syndromes, familial glioblastoma is thought to be multifactorial and autosomal recessive.^31–33^ Genome-wide association studies (GWAS) have identified several risk loci for glioblastoma, but causality of specific variants or genes in these regions remains unclear.^24, 34^

Taken together, in sporadic and/or late onset glioblastoma cases the prevalence and contribution of pathogenic germline variants (PGVs) remains unclear. It is, therefore, of interest to systematically analyze the complete germline genome of unselected glioblastoma patients, including small and structural variants, to identify genes with PGVs as potential candidates for cancer predisposition. This study thus aimed to gain novel insight into the prevalence of genetic predisposition to glioblastoma by retrospectively analyzing germline data of an unselected, adult glioblastoma patient population.

## Materials and Methods

### Patient inclusion

For this retrospective, germline analysis study, whole genome sequencing data from the Hartwig Medical Foundation (Amsterdam, the Netherlands) database was used. All patients that contributed to this database have consented to reuse of their data, including germline data, for cancer research purposes. All adult patients (i.e. from 18 years and older) diagnosed with nervous system cancer (disease ontology ID: 3093) and whose data was stored in the database before November 1, 2023, were eligible. Patients were mainly collected in the context of the CPCT-02 (NCT01855477) and GLOW (NCT05186064) studies. Hereafter, the patient selection was further filtered based on tumor type, and only gliomas WHO grade 4 were included in final analyses (*n* = 98). Sampling in these patients was performed after recurrent disease. Family history of malignant neoplasms was not taken into account. Patient consent was based on a broad consent intending publicly available access-controlled data for academic cancer research related requests. For this study, a Data Access Request (DR-310) was signed to obtain the genome-wide germline and somatic data. All samples were de-identified and keys between study number and patient number were stored solely locally in the hospitals.

### Whole genome sequencing

All samples were sequenced at Hartwig Medical Foundation as per ISO-accredited diagnostic standards (ISO17025), as described previously.^35^ Shortly, tumor samples with at least 20% tumor purity were deep-sequenced on Illumina Novaseq 6000 to an average depth of 90-100x. The blood control samples were sequenced to a depth of 30-35x. Somatic and germline variant calling was done using the open-source Hartwig WiGiTS toolset (https://github.com/hartwigmedical/hmftools v5_33). Also, tumor heterogeneity and presence of non-tumor cells in the tumor sample were computed (https://github.com/hartwigmedical/hmftools/tree/master/purple) and accounted for. The strategy for this germline analysis has been validated previously.^36^

### Selection of relevant genes

Because of interpretation challenges and limited statistical power associated with the number of available patients compared to the vast search space of the genome, as well as the expected limited penetrance of individual genes, it was considered not feasible to perform a sufficiently powered genome-wide association study for analysis of variants that might be involved in glioblastoma predisposition. Hence, a manually curated list of known cancer-associated genes was created to first explore potential involvement of candidate genes. As a basis, the reportable germline gene list used as part of the pan-cancer routine diagnostic analysis pipeline from Hartwig was used.^36^ This gene panel is based on national guidelines^37^ and experience at the Netherlands Cancer Institute and was for this study expanded with genes from several other cancer predisposition gene panels: a germline driver catalogue previously described and curated by Priestley et al.^35^, a subset of genes from the American College of Medical Genetics and Genomics (ACMG)^38^, the Hereditary Cancer Gene Curation Expert Panel from ClinGen^39^, the adult solid tumors cancer susceptibility panel created by National Health Service (NHS) and Genomics England^40^, and from the literature.^20,30^ After comparing these different gene lists, a comprehensive list of 170 genes was generated for the current germline predisposition analysis. For all of these genes the likely mechanism of action was determined as either oncogene or tumor suppressor gene (*Supplementary Table 1*).

### Small variant calling

Small variants include stop-gain mutations, frameshifts due to small insertions or deletions, inframe deletions, inframe insertions, missense mutations and splice site mutations. Within the standard pipeline workflow of Hartwig (https://github.com/hartwigmedical/pipeline5), small variants in both tumor and germline are called by the algorithm ‘Somatic Alterations in Genome’ (SAGE; v3.2) (https://github.com/hartwigmedical/hmftools/tree/master/sage). SAGE is a precise and highly sensitive caller for single nucleotide variants (SNVs), multiple nucleotide variants ≤32 base pairs (MNVs) and small insertions and deletions (InDels). In the standard data processing workflow of Hartwig, SAGE is given a panel containing the regions of genes of interest for germline analysis in a Browser Extensible Data (BED) format (*Supplementary Table 1*). For our selected gene panel, a custom BED file (https://github.com/MvOglow/germlineGBM.git) was created using the in-house tool HMF Gene Utilities (v1.1, https://github.com/hartwigmedical/hmftools/tree/master/gene-utils) which used the GENCODE coordinates for the Genome Reference Consortium Human Build 37 (GRCh37) definitions. All raw compressed reference-oriented alignment map (CRAM) files containing the aligned sequencing reads for the included patients were re-processed with SAGE using the default germline run parameters (v3.4; *Supplementary Figure 1*) and these custom gene regions. Subsequently, this data was annotated and filtered using ‘Prediction and Annotation of Variant Effects’ (PAVE) germline (v1.6) (https://github.com/hartwigmedical/hmftools/tree/master/pave) using the default germline parameters (*Supplementary Figure 2*).

Hereafter, variants annotated as having only synonymous canonical coding effects were removed from the output files. To reduce inclusion of common neutral population variants and potential false positives, additional filters were used next to the default SAGE filters: (1) variants with a Genome Aggregation Database (gnomAD; v2.1.1^41^) population frequency >1% were removed and classified as population variance; (2) germline variants with a low recalibrated quality score (see below) were removed and (3) germline variants with a frequency ≥5% in the Hartwig database (*n* = 5,778, excluding the patients included in this study) were removed as these are likely population variants specific to the Dutch population. SAGE accounted for false positive calls or poor sensitivity by recalibrating the empirical base quality score provided by the sequencer. The ad-hoc cut-off based on these recalibrated Phred-scaled quality scores was determined using a density plot of the recalibrated Phred quality of all obtained variants for the included patients and set at 235.6 for variants to be included in further analyses (*Supplementary Figure 3*).

### Structural variants and copy number variations calling

By default, structural variants (SVs) and copy number variations (CNVs) were called genome-wide by GRIDDS2 in the Hartwig pipeline.^42^ After processing, this data was annotated and filtered by GRIPPS germline and stored in a SQL database (pipeline release v5.33). All SVs and CNVs within the regions defined in the BED file were obtained from the SQL database. Because gnomAD does not provide population frequencies for SVs, the data was filtered based on the variant frequency within the Hartwig database (excluding the patients included in this study). All obtained SVs occurring in ≥5% of all other patients in the Hartwig database were excluded. Since the Hartwig databases contained 5,778 patients next to the patients included in this study, SVs occurring in ≥289 patients were discarded.

The interpretability of copy number gains is low as there is no international consensus on the significance of differences between the exact number of copies, e.g. three versus more than three copies. Moreover, most genes are tumor suppressor genes. Therefore, we assessed only copy number losses and no copy number gains.

### Clinical significance

The clinical significance of variants was based on their annotation in ClinVar, a public archive of human genetic variants and interpretations of their significance to disease.^43^ The main conclusions in our study were based on ‘pathogenic’ and ‘likely pathogenic’ variants. Variants of unknown significance (VUS) were not studied. To direct the potential effect of these variants on the functional protein, the Ensembl Variant Effect Predictor (VEP) was used.^44^ All shortlisted variants were manually reviewed by a clinical laboratory geneticist (RK) to determine pathogenicity according to routine diagnostic procedures, all likely-pathogenic (class 4) and pathogenic variants (class 5) were considered as pathogenic germline variants (PGVs). As a second step to assess the clinical significance of PGVs, tumor type-specific manual curation and tumor genome analysis was performed. The following subdivision was used: category 1 were causal events (gene associated with glioblastoma + matching tumor findings), category 2 were known predisposition genes but without demonstrated causality (gene associated with glioblastoma without matching tumor findings, or gene not associated but having matching tumor findings), and category 3 contained variants less likely to contribute to glioblastoma.

### Tumor sample analysis

For tumor suppressor genes, the common model for pathogenicity is that both alleles of the gene become inactivated in the tumor. In case of germline predisposition, the second allele is typically inactivated by a second mutation or loss of heterozygosity (LOH, although epi-genetic inactivation through methylation is also possible). Therefore, we assessed all candidate genes for somatic events, and, in case of LOH, determined if the normal or mutated germline allele was lost. In addition, we explored if any of the candidate genes was also a common somatic driver in glioblastoma patients, i.e. inactivated bi-allelic by somatic events. Finally, mutational signatures were studied for DNA repair genes. In case of splice site variants, RNA sequencing data (which was available for approximately 80% of the patients) was used to validate the impact of the variant at transcript levels. A graphic overview of the methods for identification of predisposition can be found in *Figure 1*.

**Figure 1.**
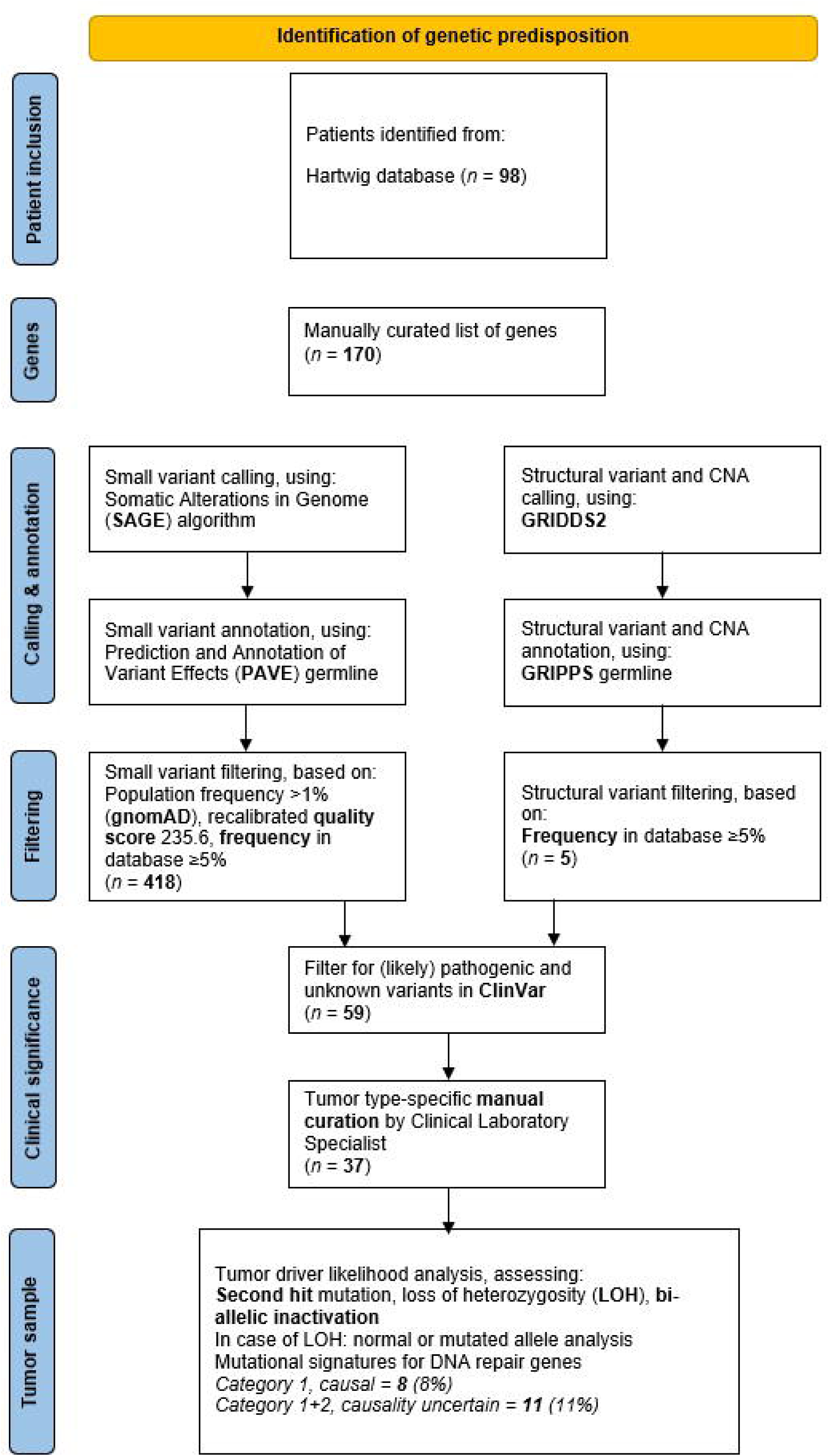
Flowchart methods

### Statistics

Sociodemographic characteristics were compared by using chi-square test for categorical variables and t-test for continuous variables. In case of violation of the normality assumption, a non-parametric test was used for the continuous variables.

## Results

### Patient characteristics

A total of 98 patients met the inclusion criterium of ‘adult glioma WHO grade 4’ and were included in this study for germline predisposition analysis. Of these, 70.4% (69/98) were male and the median age for males and females was 61 years. Most of the patients had a primary, isocitrate dehydrogenase (*IDH*) wildtype glioblastoma (93.9%, 92/98) while 6.1% (6/98) of the tumors had a somatic *IDH* mutation (classifying them as astrocytoma WHO grade 4). The family history, particularly regarding the occurrence of malignant neoplasms, was unidentified.

### Germline findings in an average glioblastoma population

After filtering for canonical coding effects, gnomAD population frequency and quality score, a total of 418 small variants and five structural variants (SVs) were detected in 107 of the 170 different genes. Filtering for variants that were annotated as ‘(likely) pathogenic’ or ‘unknown’ in ClinVar following manual curation, resulted in a total of 30 (including three SVs) PGVs in 18 different genes in 25 unique patients (25.5% of all patients). Of these 30 PGVs, 11 were observed in genes with an explicitly recessive inheritance and 19 in genes having dominant inheritance. All 11 PGVs in recessive genes were monoallelic and, therefore, excluded from overall prevalence, because only biallelic or compound heterozygous germline variants in such genes are considered as having associated hereditary risks (*Table 1*).

**Table 1.**
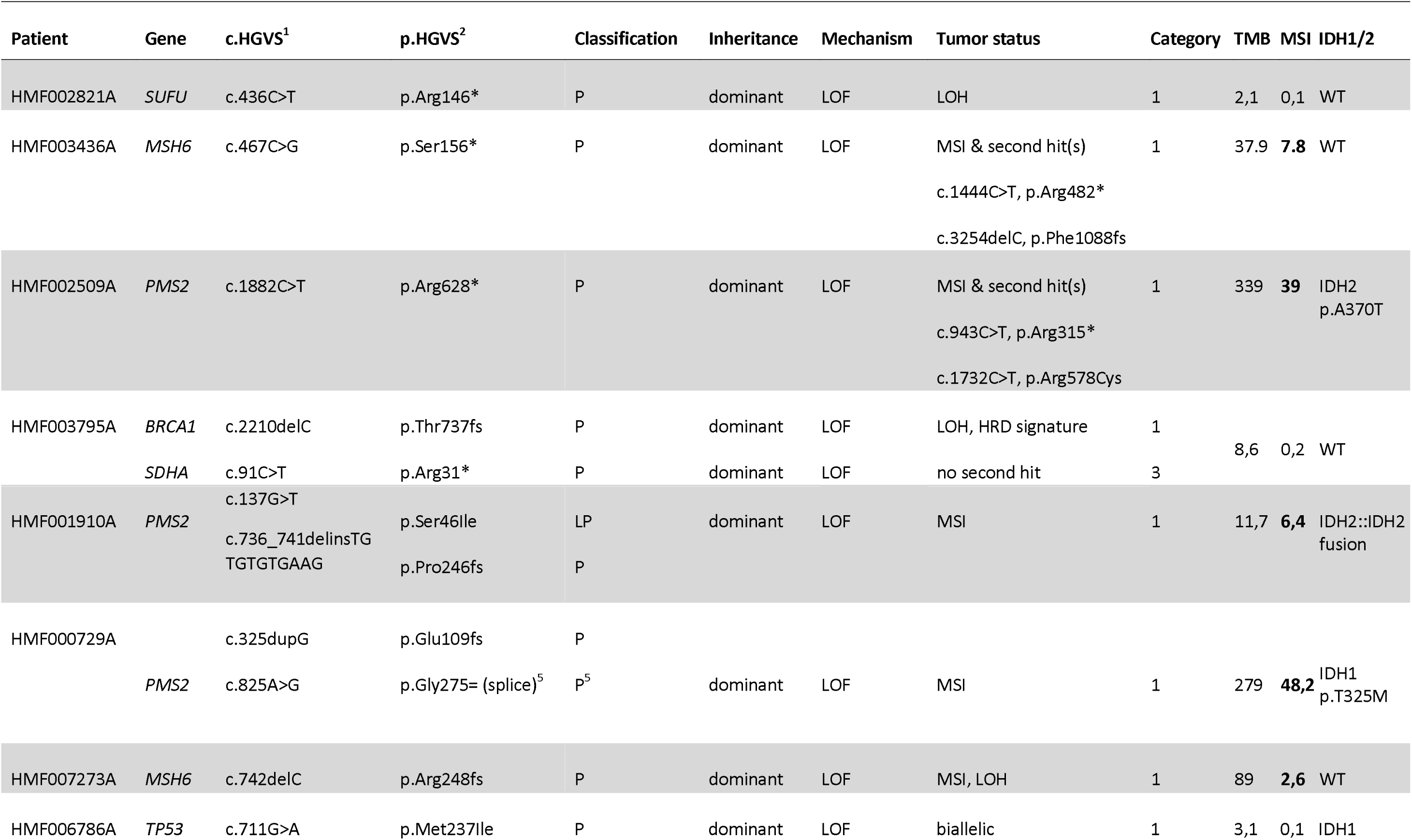
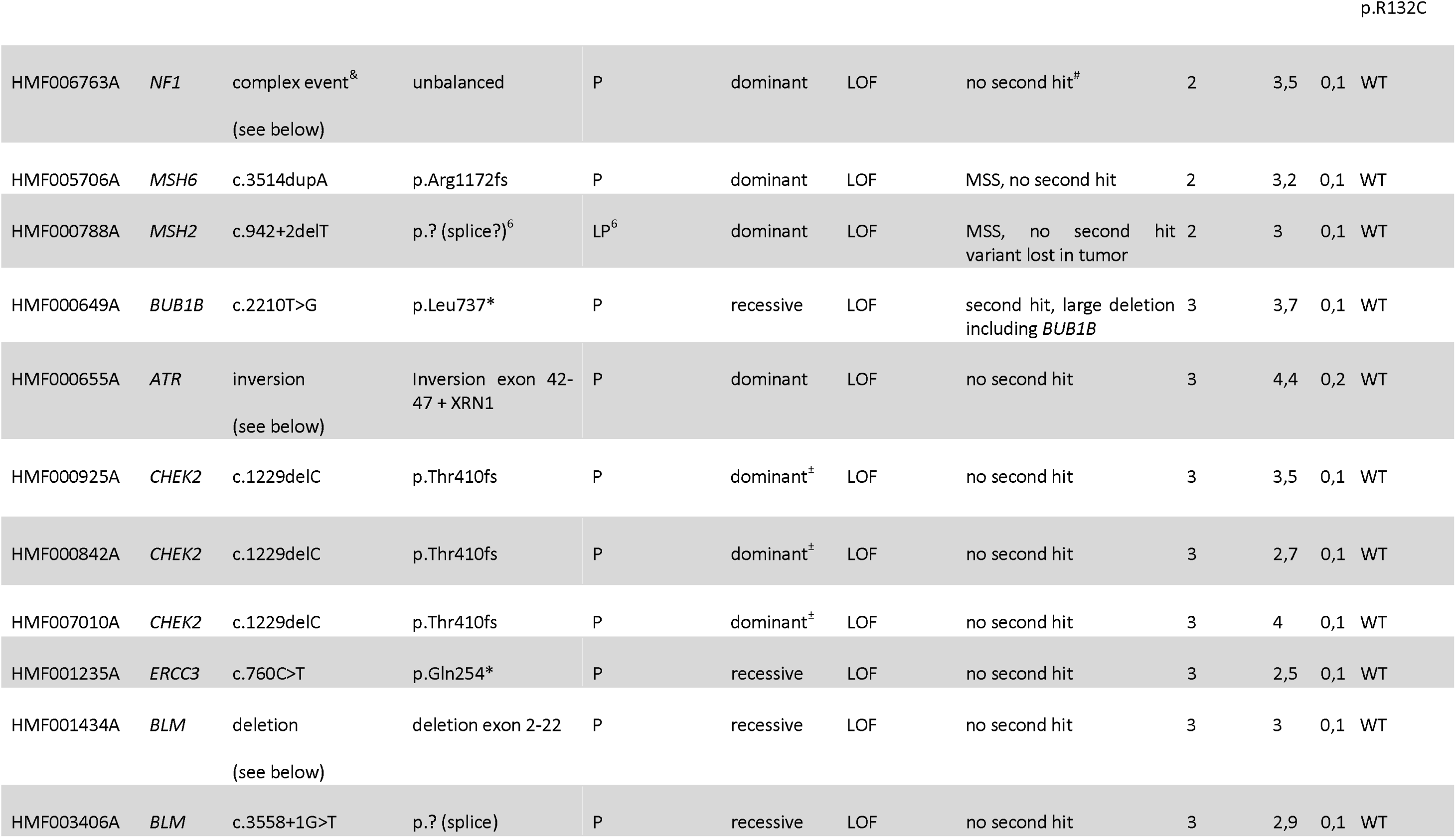
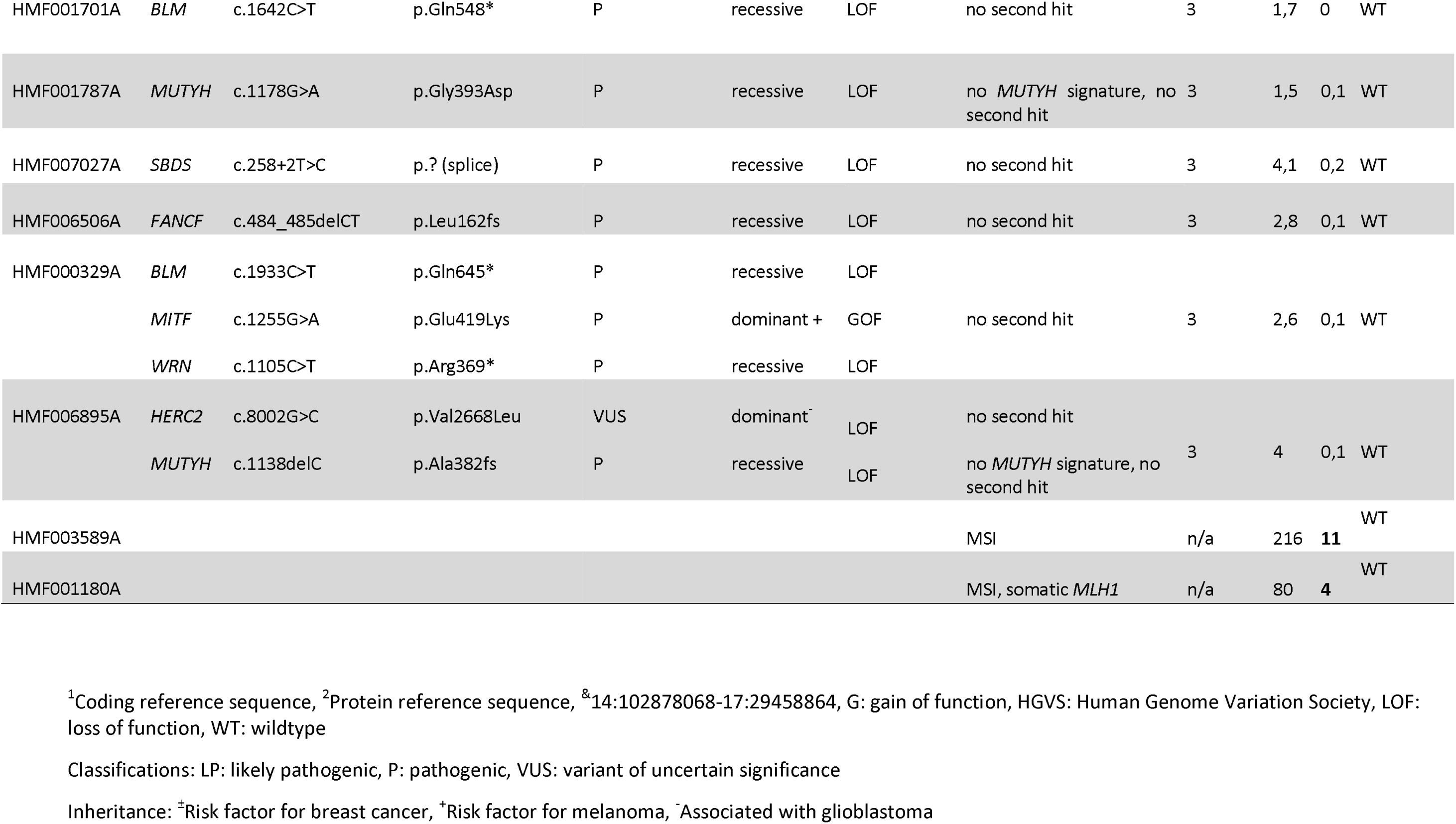
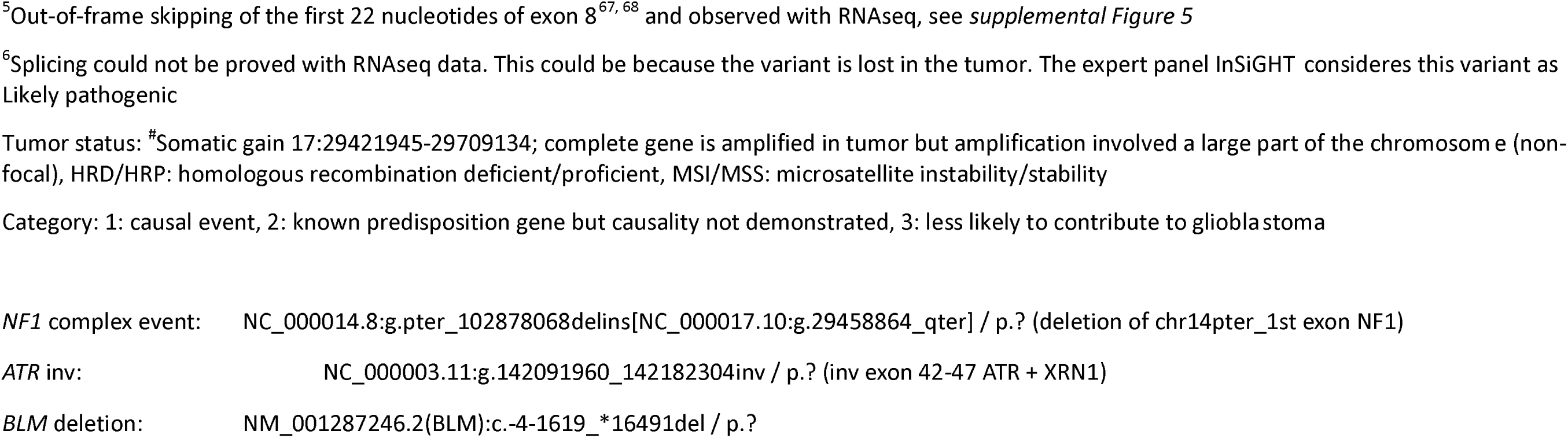
Findings after manual curation of possibly interesting variants.

The 19 dominant inheritance PGVs were present in 11 different genes in 16 unique patients (16% of all patients). Six of these PGVs were in cancer predisposition genes (*ATR*, *CHEK2* (3x), *SDHA* and *MITF*) without an established association with familial glioblastoma. Interestingly, the majority, 13 PGVs in 11 patients, were in established cancer predisposition genes with a strong association with familial glioblastoma (*MSH6* (3x), *PMS2* (5x), *MSH2*, *TP53*, *NF1* and *BRCA1*) or with medulloblastoma (*SUFU*). Thus, the prevalence of known genetic predisposition to glioblastoma was 11% (11/98) in our unselected cohort, with additional candidates in another 5.1% of patients (5/98).

### Genetic predisposition driving glioblastoma oncogenesis

As most predisposition genes involve tumor suppressors, all candidate causal events were assessed for second hit (somatic) events in the tumor data. PGVs with a second (somatic) event are considered causal for glioblastoma oncogenesis. For all six PGVs (*ATR*, *CHEK2* (3x), *SDHA* and *MITF*) without an established association with familial glioblastoma and for 10 out of 11 PGVs in recessive genes (*BLM* (4x), *ERCC3*, *MUTYH* (2x), *FANCF*, *SBDS* and *WRN*), no second (somatic) event (small variant or structural variant resulting in LOH) or matching mutational signature was detected in the tumor. Thus, those variants, except for possibly *BUB1B*, were unlikely to contribute to the development of glioblastoma in our cohort (category 3 – see *Table 1*). Additionally, for three patients with PGVs in genes with a strong association with familial glioblastoma (*NF1*, *MSH6* and *MSH2*) also no second (somatic) event or expected matching mutational signature was detected, indicating that for these variants the causality for tumorigenesis in these patients remains unclear (category 2 – see *Table 1*).

Importantly, for the remaining 10 PGVs that were identified in genes with a strong association with familial glioblastoma or medulloblastoma (*SUFU*, *MSH6* (2x), *PMS2* (5x), *TP53* and *BRCA1*), a second (somatic) event and/or a matching mutational signature was identified in the tumor. These variants were present in eight different patients, resulting in a proven germline predisposition rate of 73% in the patients with relevant PGVs (8/11). Of interest, two of these patients most likely have constitutional mismatch repair deficiency (CMMRD) syndrome, since they each harbored two PGVs in *PMS2* and both were microsatellite instable with a high tumor mutational burden (*Table 1*).

### DNA damage response – significant role for mismatch repair (MMR) in glioblastoma

The known pathogenic predisposition variants in 11 patients could be divided in two main mechanisms. First, two patients had PGVs in genes involved in cell proliferation/survival (Ras/mitogen-activated protein kinase pathway; NF1 & Shh signaling pathway; *SUFU*). Second, nine patients had 11 PGVs in genes involved in the DNA damage response or cell cycle pathway (*TP53*, *BRCA1*, *PMS2*, *MSH6* and *MSH2*). These included a patient showing LOH for *TP53* and another patient showing LOH for *BRCA1* along with a homologous recombination deficiency (HRD) footprint. The tumor in this patient underwent whole genome duplication after LOH (*Supplement Figure 4)*.

The majority of patients was thus found to harbor a PGV in one of the mismatch repair (MMR: MSH2, PMS2, MLH1, MSH6) genes (7 out of 11). By measuring microsatellite instability (MSI) based on WGS, we observed that, within the total cohort, seven patients had > 1.3 microsatellite Indels Per Mb (overall average 1.3, median 0.12) and six of seven patients had ≥ 4 microsatellite Indels Per Mb (diagnostic cutoff of WGS handled by Hartwig Medical Foundation – see *Figure 2A*). For one of the seven patients with MSI no evidence for either germline or somatic mutations in any of the four MMR genes was found. For the remaining six patients with MSI, one patient with somatic loss of function of *MLH1* and five patients with germline loss of function of *MSH6* (2x) or *PMS2* (3x) matched with MSI (*Figure 2A*).

**Figure 2.**
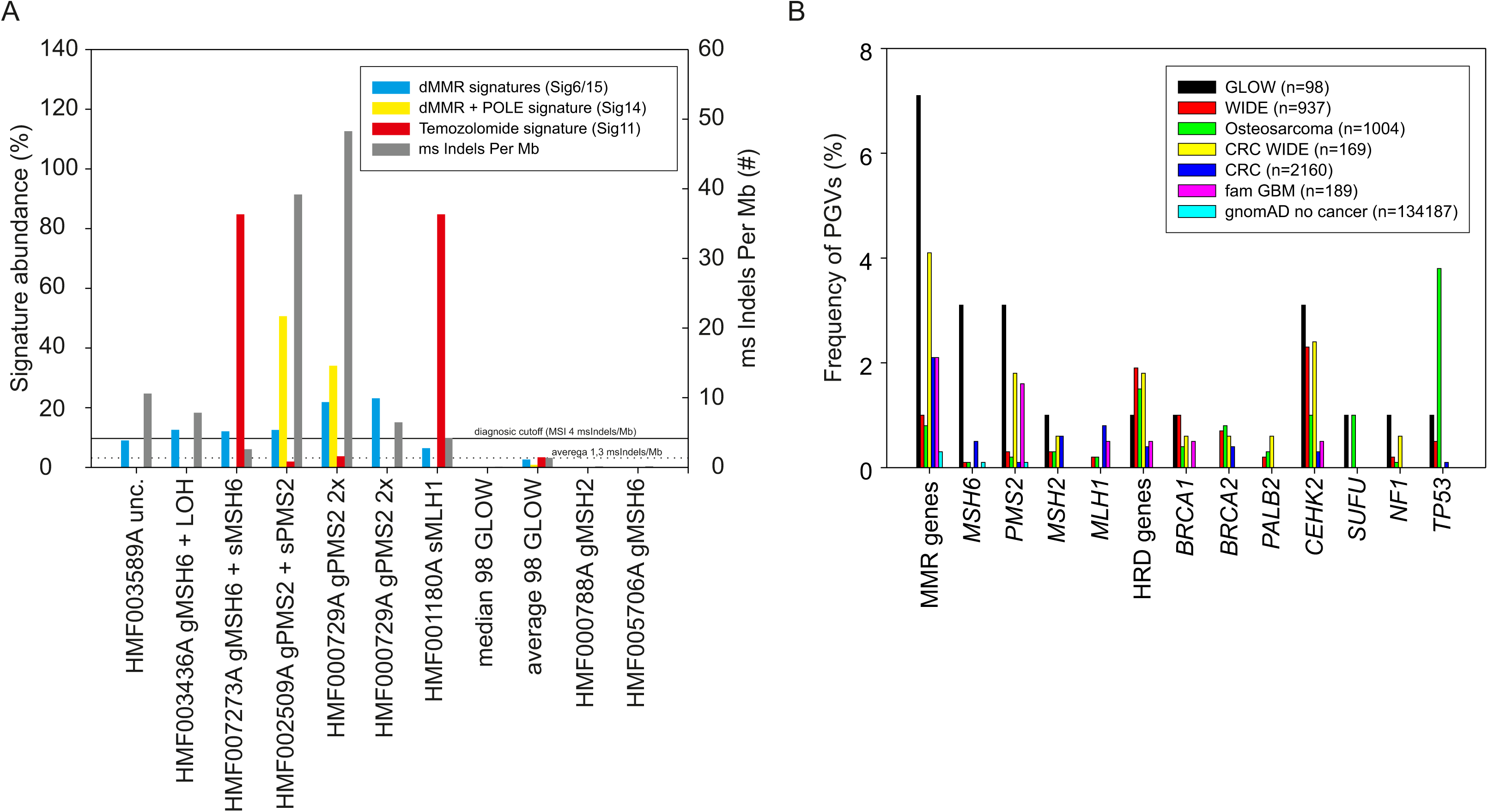
Significant role for mismatch repair (MMR) in glioblastoma. (A) Cosmic single base substitution (SBS) signatures for dMMR (Sig 6+15) dMMR+POLE (Sig14) and Temozolomide (Sig11) and number of ms Indels per Mb are depicted for seven patients within the total cohort having > 1.3 microsatellite Indels Per Mb. (B) Frequency of pathogenic germline variants (PGVs) in the genes as described in the GLOW study versus other cohorts. CRC: colorectal cancer^46^, CRC WIDE: subgroup WIDE colorectal cancer patients^36^, fam GBM: familial glioblastoma cohort^20^, GLOW: current composite cohort, gnomad: non-cancer reference cohort^41^, HRD genes: homologous recombination deficiency genes (*BRCA1/2* & *PALB2*), dMMR genes: deficient mismatch repair genes (*MSH6*, *PMS2*, *MSH2* & *MLH1*), MSI: microsatellite instability, osteosarcoma^45^, WIDE: metastatic cancer^36^

The percentage of patients with PGVs in MMR genes within this unselected glioblastoma cohort were compared to the percentage of patients with PGVs in MMR genes within other unselected cancer cohorts^20, 36, 45, 46^ and the gnomAD v2.11 (non-cancer) cohort^41^. Although numbers remain small, a higher than expected frequency of patients with glioblastoma carrying a PGV in MMR genes was seen, with the biggest difference for *MSH6* and *PMS2* (*Figure 2B*).

## Discussion

This study showed the germline predisposition in a cohort of 98 adult glioblastoma patients. In 11% of the patients, pathogenic germline variants (PGVs) were observed in genes previously associated with familial glioblastoma; thus these PGVs likely contributed to the oncogenesis of these unselected glioblastoma patients. PGVs were found in the following genes: *BRCA1*, *MSH6*, *PMS2, TP53, NF1* and *SUFU*. Furthermore, for ten PGVs in *SUFU*, *MSH6* (2x), *PMS2* (5x), *TP53* and *BRCA1*, in eight different patients causality was proven, since second (somatic) events and/or matching mutational signature were detected. Several of these PGVs were in predisposition genes that are increasingly important for (targeted) therapy selection and for all findings counseling by a clinical geneticist is indicated. Mismatch repair deficiency formed the main mechanism of the unselected cohort, with 7.1% of the patients harboring a PGV in one of the mismatch repair (MMR) genes, including five patients with microsatellite instability.

The results of this study are unique in several aspects. First, no preselection based on personal and/or family history of malignant neoplasms was applied to the study cohort. Second, the pairing of both blood and tumor tissue samples allowed for verification of the causality of potentially interesting events. Third, since all patients underwent paired WGS testing combined with RNA sequencing (∼80%), we were able to not only study point mutations (which is a limitation in most of the cancer predisposition research) but also copy number variations, structural variants, splice site variants (*supplementary Figure 5*) and mutational signatures.

We detected a number of PGVs in dominant and recessive genes without proven causality for glioblastoma, since the tumor sample analyses did not show second hit mutations in almost all of these cases. In the Netherlands, observed putative PGVs in dominant genes that do not match the tumor type (*ATR*, *CHEK2* (3x), *SDHA* and *MITF*) are normally not reported back to the patient, except if there is a matching personal and/or familial history. Unfortunately, in the current retrospective study design, we were not able to identify the pedigrees of the patients with PGVs, making further details of the inheritance pattern and possible consequences for family members impossible. In recessive genes, all 11 PGVs were monoallelic and considered low/no risk for cancer predisposition. Still, these variants potentially modified the genesis of the tumor as risk loci associated with susceptibility to glioblastoma. Unfortunately, our study lacked sufficient power to study these monoallelic PGVs in recessive genes in a statistically sound manner. Interestingly, in one patient with a PGV in *BUB1B*, the remaining wildtype allele was somatically lost due to a large deletion. The causality of the PGV in this recessive gene could not been demonstrated, although there is evidence for the role of *BUB1B* as a (pan-)cancer predisposing gene^47^, including glioblastoma.^48^ When variants like these are identified, they are normally not reported back to the patient. Because these variants do not have any relevance for the patient nor for the patient’s family except if there is consanguinity in the family, no genetic counselling and testing is recommended.

Currently, the international guideline of the European Association of Neuro-Oncology (EANO) on the diagnosis and treatment of diffuse gliomas of adulthood recommend genetic counselling in patients with ‘relevant germline variants or suspected hereditary cancer syndromes.’^49^ This recommendation is based on low level evidence (i.e. class IV, level C evidence) and did not specify which germline variants are considered relevant. The familial tumor syndromes associated with gliomagenesis named in this EANO guideline include neurofibromatosis type I, tuberous sclerosis, Turcot syndrome, Li-Fraumeni syndrome and Lynch syndrome. Other international guidelines of neuro-oncology or medical oncology societies lack recommendations on germline testing and genetic counseling of gliomas in adults.^50, 51^ However, the more recent EANO guideline on molecular testing of gliomas in adults recommend genetic counseling prior to germline testing, as for instance specific attention is paid to MMR gene deficiencies.^52^ Yet, most of the PGVs found in our study are currently not tested for in most of the Dutch laboratories.^53^

As the use of comprehensive tumor genetic and genomic diagnostic tests continues to grow, the detection of PGVs is occurring more frequently than previously expected.^36, 48, 54, 55^ In our unselected cohort, many PGVs are identified in genes such as *BRCA1*, *MSH6*, *PMS2*, and *NF1*, which are crucial not only for germline follow-up but also for selecting appropriate therapies, particularly immune-based or targeted treatments, as observed in other tumor types. For example, melanoma, MMR deficient colorectal cancer, and other non-colorectal MMR deficient tumors have shown remarkable responses to immunotherapy.^13–16, 56^ While some glioblastoma patients exhibit long-term responses to immunotherapy, this treatment has shown limited efficacy in over 90% of unselected glioblastoma cases.^57–60^ Among those who responded (partially or fully), most likely were patients with hypermutated tumors, possibly due to MMR deficiency or MMR deficiency + *POLE* defects.^57, 61–65^ Our findings indicate that most of these hypermutated tumors harbor a PGV in one of the MMR genes. Thus, comprehensive tumor genetic and genomic profiling for glioblastoma patients requires an integrated approach that facilitates appropriate referral to clinical geneticists.

This study has some limitations that have to be considered. First, this type of research cannot be done without making assumptions. Assumptions were not only made when defining the pathogenicity of variants^38^, but essentially every single step in our methods, e.g. variant calling, annotation, filtering, curation involved choices based on assumptions. Although these are based on generally accepted international standards, changes over time based on progressive insights may impact outcomes. A second limitation is the relatively small sample size, which hampered statistically powered analyses of the PGVs. Third, our cohort contained six patients (6.1%) with a somatic *IDH* mutation, which might be extra relevant, in terms of prognostic relevance, in the context of MMR deficiency.^66^ Lastly, due to consent and privacy regulation limitations, we were not able to assess the pedigrees of the patients with PGVs, making assessment of the inheritance pattern and possible consequences for family members impossible.

To conclude, this study investigated the germline predisposition to glioblastoma in an average adult glioblastoma population. 11% of these patients had a pathogenic germline variant that (likely) predisposed to the development of the glioblastoma, with potential associated therapy options. The results could guide clinicians who have to inform patients about broad molecular tests for personalized medicine and its associated putative germline findings, once current gene panels are adapted to these findings.

## Data Availability

The underlying research are partly facilitated by Hartwig Medical Foundation and the Center for Personalized Cancer Treatment (CPCT) which have generated, analysed and made available data for this research. Data can be requested via https://www.hartwigmedicalfoundation.nl/en/data/data-access-request/. Hartwig Medical Foundation is willing to share with external qualified researchers access to patient-level data and supporting clinical documents. These requests are reviewed and approved by an independent review committee on the basis of scientific merit. All data provided are anonymized to respect the privacy of patients who have participated in the study, in line with applicable laws and regulations.

## Authorship

Study concept and design: M.P.v.O, D.R.v.V., M.L.D.B., E.C. Material preparation, data collection, and analysis: M.P.v.O., D.R.v.V., J.d.L., E.C., R.K. Writing and revision of manuscript: M.P.v.O., D.R.v.V., J.d.L., F.Y.F.d.V., M.L.D.B., E.C., R.K.

## Supplementary

**Supplementary Figure 1** Example of command-line arguments that were used to run SAGE in default germline mode

**Supplementary Figure 2** Example of command line arguments that were used to run PAVE in default germline mode

**Supplementary Figure 3** Quality density plot of small variant scores

The SAGE recalibrated Phred quality score was used to create a density plot. The elbow of the graph at recalibrated Phred quality score = 235.6 was used as minimum cut-off while filtering the patients with observed small variants

**Supplementary Figure 4** Circos plot (left) and main SBS Cosmic mutational signatures detected (right) of HMF006786A.

**Supplementary Figure 5** Insilicopredictions for PMS2:c.825A>G (p.Gly275=) and RNAseqanalysis of PMS2 exon 1-12 forHMF000729A (PMS2:c.825A>G & c.325dup)

**Supplementary Table 1** Overview of the genes included in the gene panel that was used

See *Methods* section for composition of the gene panel. ONCO: proto-onco gene; TSG: tumor suppressor gene

## References

1. Stupp R, Mason WP, van den Bent MJ, Weller M, Fisher B, Taphoorn MJ, et al. Radiotherapy plus concomitant and adjuvant temozolomide for glioblastoma. N Engl J Med. 2005;352(10):987–96.

2. Stupp R, Hegi ME, Mason WP, van den Bent MJ, Taphoorn MJ, Janzer RC, et al. Effects of radiotherapy with concomitant and adjuvant temozolomide versus radiotherapy alone on survival in glioblastoma in a randomised phase III study: 5-year analysis of the EORTC-NCIC trial. Lancet Oncol. 2009;10(5):459–66.

3. Akgül S, Patch AM, D’Souza RCJ, Mukhopadhyay P, Nones K, Kempe S, et al. Intratumoural Heterogeneity Underlies Distinct Therapy Responses and Treatment Resistance in Glioblastoma. Cancers (Basel). 2019;11(2).

4. van de Geer WS, Hoogstrate Y, Draaisma K, Robe PA, Bins S, Mathijssen RHJ, et al. Landscape of driver gene events, biomarkers, and druggable targets identified by whole-genome sequencing of glioblastomas. Neurooncol Adv. 2022;4(1):vdab177.

5. Patel AP, Tirosh I, Trombetta JJ, Shalek AK, Gillespie SM, Wakimoto H, et al. Single-cell RNA-seq highlights intratumoral heterogeneity in primary glioblastoma. Science. 2014;344(6190):1396–401.

6. Khalafallah AM, Huq S, Jimenez AE, Serra R, Bettegowda C, Mukherjee D. "Zooming in" on Glioblastoma: Understanding Tumor Heterogeneity and its Clinical Implications in the Era of Single-Cell Ribonucleic Acid Sequencing. Neurosurgery. 2021;88(3):477–86.

7. Roepman P, de Bruijn E, van Lieshout S, Schoenmaker L, Boelens MC, Dubbink HJ, et al. Clinical Validation of Whole Genome Sequencing for Cancer Diagnostics. J Mol Diagn. 2021;23(7):816–33.

8. Samsom KG, Schipper LJ, Roepman P, Bosch LJ, Lalezari F, Klompenhouwer EG, et al. Feasibility of whole-genome sequencing-based tumor diagnostics in routine pathology practice. J Pathol. 2022;258(2):179–88.

9. van Opijnen MP, Broekman MLD, de Vos FYF, Cuppen E, van der Hoeven JJM, van Linde ME, et al. Study protocol of the GLOW study: maximising treatment options for recurrent glioblastoma patients by whole genome sequencing-based diagnostics-a prospective multicenter cohort study. BMC Med Genomics. 2022;15(1):233.

10. Castro E, Romero-Laorden N, Del Pozo A, Lozano R, Medina A, Puente J, et al. PROREPAIR-B: A Prospective Cohort Study of the Impact of Germline DNA Repair Mutations on the Outcomes of Patients With Metastatic Castration-Resistant Prostate Cancer. J Clin Oncol. 2019;37(6):490–503.

11. Tutt ANJ, Garber JE, Kaufman B, Viale G, Fumagalli D, Rastogi P, et al. Adjuvant Olaparib for Patients with BRCA1- or BRCA2-Mutated Breast Cancer. N Engl J Med. 2021;384(25):2394–405.

12. Litton JK, Rugo HS, Ettl J, Hurvitz SA, Gonçalves A, Lee KH, et al. Talazoparib in Patients with Advanced Breast Cancer and a Germline BRCA Mutation. N Engl J Med. 2018;379(8):753–63.

13. Harrold EC, Foote MB, Rousseau B, Walch H, Kemel Y, Richards AL, et al. Neoplasia risk in patients with Lynch syndrome treated with immune checkpoint blockade. Nat Med. 2023;29(10):2458–63.

14. de Gooyer PGM, Verschoor YL, van den Dungen LDW, Balduzzi S, Marsman HA, Geukes Foppen MH, et al. Neoadjuvant nivolumab and relatlimab in locally advanced MMR-deficient colon cancer: a phase 2 trial. Nat Med. 2024;30(11):3284–90.

15. Marabelle A, Le DT, Ascierto PA, Di Giacomo AM, De Jesus-Acosta A, Delord JP, et al. Efficacy of Pembrolizumab in Patients With Noncolorectal High Microsatellite Instability/Mismatch Repair-Deficient Cancer: Results From the Phase II KEYNOTE-158 Study. J Clin Oncol. 2020;38(1):1–10.

16. Yu JH, Xiao BY, Tang JH, Li DD, Wang F, Ding Y, et al. Efficacy of PD-1 inhibitors for colorectal cancer and polyps in Lynch syndrome patients. Eur J Cancer. 2023;192:113253.

17. Mahdavi M, Nassiri M, Kooshyar MM, Vakili-Azghandi M, Avan A, Sandry R, et al. Hereditary breast cancer; Genetic penetrance and current status with BRCA. J Cell Physiol. 2019;234(5):5741–50.

18. Jasperson KW, Tuohy TM, Neklason DW, Burt RW. Hereditary and familial colon cancer. Gastroenterology. 2010;138(6):2044–58.

19. Gestrich CK, Jajosky AN, Elliott R, Stearns D, Sadri N, Cohen ML, Couce ME. Molecular Profiling of Pediatric and Adult Glioblastoma. Am J Clin Pathol. 2021;155(4):606–14.

20. Choi DJ, Armstrong G, Lozzi B, Vijayaraghavan P, Plon SE, Wong TC, et al. The genomic landscape of familial glioma. Sci Adv. 2023;9(17):eade2675.

21. Wrensch M, Lee M, Miike R, Newman B, Barger G, Davis R, et al. Familial and personal medical history of cancer and nervous system conditions among adults with glioma and controls. Am J Epidemiol. 1997;145(7):581–93.

22. Hemminki K, Tretli S, Sundquist J, Johannesen TB, Granström C. Familial risks in nervous-system tumours: a histology-specific analysis from Sweden and Norway. Lancet Oncol. 2009;10(5):481–8.

23. Sadetzki S, Bruchim R, Oberman B, Armstrong GN, Lau CC, Claus EB, et al. Description of selected characteristics of familial glioma patients - results from the Gliogene Consortium. Eur J Cancer. 2013;49(6):1335–45.

24. Shete S, Lau CC, Houlston RS, Claus EB, Barnholtz-Sloan J, Lai R, et al. Genome-wide high-density SNP linkage search for glioma susceptibility loci: results from the Gliogene Consortium. Cancer Res. 2011;71(24):7568–75.

25. Rice T, Lachance DH, Molinaro AM, Eckel-Passow JE, Walsh KM, Barnholtz-Sloan J, et al. Understanding inherited genetic risk of adult glioma - a review. Neurooncol Pract. 2016;3(1):10–6.

26. Ostrom QT, Bauchet L, Davis FG, Deltour I, Fisher JL, Langer CE, et al. The epidemiology of glioma in adults: a "state of the science" review. Neuro Oncol. 2014;16(7):896–913.

27. Ryan NAJ, Glaire MA, Blake D, Cabrera-Dandy M, Evans DG, Crosbie EJ. The proportion of endometrial cancers associated with Lynch syndrome: a systematic review of the literature and meta-analysis. Genet Med. 2019;21(10):2167–80.

28. Bainbridge MN, Armstrong GN, Gramatges MM, Bertuch AA, Jhangiani SN, Doddapaneni H, et al. Germline mutations in shelterin complex genes are associated with familial glioma. J Natl Cancer Inst. 2015;107(1):384.

29. McDonald MF, Prather LL, Helfer CR, Ludmir EB, Echeverria AE, Yust-Katz S, et al. Prevalence of pathogenic germline variants in adult-type diffuse glioma. Neurooncol Pract. 2023;10(5):482–90.

30. Jacobs DI, Fukumura K, Bainbridge MN, Armstrong GN, Tsavachidis S, Gu X, et al. Elucidating the molecular pathogenesis of glioma: integrated germline and somatic profiling of a familial glioma case series. Neuro Oncol. 2018;20(12):1625–33.

31. Malmer B, Iselius L, Holmberg E, Collins A, Henriksson R, Grönberg H. Genetic epidemiology of glioma. Br J Cancer. 2001;84(3):429–34.

32. de Andrade M, Barnholtz JS, Amos CI, Adatto P, Spencer C, Bondy ML. Segregation analysis of cancer in families of glioma patients. Genet Epidemiol. 2001;20(2):258–70.

33. Goodenberger ML, Jenkins RB. Genetics of adult glioma. Cancer Genet. 2012;205(12):613–21.

34. Melin BS, Barnholtz-Sloan JS, Wrensch MR, Johansen C, Il’yasova D, Kinnersley B, et al. Genome-wide association study of glioma subtypes identifies specific differences in genetic susceptibility to glioblastoma and non-glioblastoma tumors. Nat Genet. 2017;49(5):789–94.

35. Priestley P, Baber J, Lolkema MP, Steeghs N, de Bruijn E, Shale C, et al. Pan-cancer whole-genome analyses of metastatic solid tumours. Nature. 2019;575(7781):210–6.

36. Koster R, Schipper LJ, Giesbertz NAA, van Beek D, Mendeville M, Samsom KG, et al. Impact of genetic counseling strategy on diagnostic yield and workload for genome-sequencing-based tumor diagnostics. Genet Med. 2024;26(2):101032.

37. Vereniging Klinische Genetica Nederland (VKGN). Tabel 3: Leidraad voor verwijzing na DNA-onderzoek in (tumor)weefsel (versie januari 2024). Available at https://artsengenetica.nl/sites/default/files/tabel-3-versie-januari-2024.pdf. Accessed 01-07-2024.

38. Miller DT, Lee K, Abul-Husn NS, Amendola LM, Brothers K, Chung WK, et al. ACMG SF v3.2 list for reporting of secondary findings in clinical exome and genome sequencing: A policy statement of the American College of Medical Genetics and Genomics (ACMG). Genet Med. 2023;25(8):100866.

39. Ritter DI, Rao S, Kulkarni S, Madhavan S, Offit K, Plon SE. A case for expert curation: an overview of cancer curation in the Clinical Genome Resource (ClinGen). Cold Spring Harb Mol Case Stud. 2019;5(5).

40. Stark Z, Foulger RE, Williams E, Thompson BA, Patel C, Lunke S, et al. Scaling national and international improvement in virtual gene panel curation via a collaborative approach to discordance resolution. Am J Hum Genet. 2021;108(9):1551–7.

41. Karczewski KJ, Francioli LC, Tiao G, Cummings BB, Alföldi J, Wang Q, et al. The mutational constraint spectrum quantified from variation in 141,456 humans. Nature. 2020;581(7809):434–43.

42. Cameron DL, Baber J, Shale C, Valle-Inclan JE, Besselink N, van Hoeck A, et al. GRIDSS2: comprehensive characterisation of somatic structural variation using single breakend variants and structural variant phasing. Genome Biol. 2021;22(1):202.

43. Landrum MJ, Lee JM, Benson M, Brown GR, Chao C, Chitipiralla S, et al. ClinVar: improving access to variant interpretations and supporting evidence. Nucleic Acids Res. 2018;46(D1):D1062–d7.

44. McLaren W, Gil L, Hunt SE, Riat HS, Ritchie GR, Thormann A, et al. The Ensembl Variant Effect Predictor. Genome Biol. 2016;17(1):122.

45. Mirabello L, Zhu B, Koster R, Karlins E, Dean M, Yeager M, et al. Frequency of Pathogenic Germline Variants in Cancer-Susceptibility Genes in Patients With Osteosarcoma. JAMA Oncol. 2020;6(5):724–34.

46. Liao H, Cai S, Bai Y, Zhang B, Sheng Y, Tong S, et al. Prevalence and spectrum of germline cancer susceptibility gene variants and somatic second hits in colorectal cancer. Am J Cancer Res. 2021;11(11):5571–80.

47. Silva MP, Ferreira LT, Brás NF, Torres L, Brandão A, Pinheiro M, et al. BUB1B monoallelic germline variants contribute to prostate cancer predisposition by triggering chromosomal instability. J Biomed Sci. 2024;31(1):74.

48. Huang KL, Mashl RJ, Wu Y, Ritter DI, Wang J, Oh C, et al. Pathogenic Germline Variants in 10,389 Adult Cancers. Cell. 2018;173(2):355–70.e14.

49. Weller M, van den Bent M, Preusser M, Le Rhun E, Tonn JC, Minniti G, et al. EANO guidelines on the diagnosis and treatment of diffuse gliomas of adulthood. Nat Rev Clin Oncol. 2021;18(3):170–86.

50. Mohile NA, Messersmith H, Gatson NT, Hottinger AF, Lassman A, Morton J, et al. Therapy for Diffuse Astrocytic and Oligodendroglial Tumors in Adults: ASCO-SNO Guideline. J Clin Oncol. 2022;40(4):403–26.

51. Stupp R, Brada M, van den Bent MJ, Tonn JC, Pentheroudakis G. High-grade glioma: ESMO Clinical Practice Guidelines for diagnosis, treatment and follow-up. Ann Oncol. 2014;25 Suppl 3:iii93–101.

52. Capper D, Reifenberger G, French PJ, Schweizer L, Weller M, Touat M, et al. EANO guideline on rational molecular testing of gliomas, glioneuronal, and neuronal tumors in adults for targeted therapy selection. Neuro Oncol. 2023;25(5):813–26.

53. van Opijnen MP, Broekman MLD, Cuppen E, Dubbink HJ, Ter Elst A, van Eijk R, et al. Next generation sequencing of high-grade adult-type diffuse glioma in the Netherlands: interlaboratory variation in the primary diagnostic and recurrent setting. J Neurooncol. 2024;166(3):485–92.

54. Zhang J, Walsh MF, Wu G, Edmonson MN, Gruber TA, Easton J, et al. Germline Mutations in Predisposition Genes in Pediatric Cancer. N Engl J Med. 2015;373(24):2336–46.

55. Schrader KA, Cheng DT, Joseph V, Prasad M, Walsh M, Zehir A, et al. Germline Variants in Targeted Tumor Sequencing Using Matched Normal DNA. JAMA Oncol. 2016;2(1):104–11.

56. Blank CU, Lucas MW, Scolyer RA, van de Wiel BA, Menzies AM, Lopez-Yurda M, et al. Neoadjuvant Nivolumab and Ipilimumab in Resectable Stage III Melanoma. N Engl J Med. 2024;391(18):1696–708.

57. Bartkowiak T, Brockman AA, Mobley BC, Harmsen H, Moots P, Merrell R, et al. Pembrolizumab alters the tumor immune landscape in a patient with dMMR glioblastoma. medRxiv. 2023.

58. Arrieta VA, Dmello C, McGrail DJ, Brat DJ, Lee-Chang C, Heimberger AB, et al. Immune checkpoint blockade in glioblastoma: from tumor heterogeneity to personalized treatment. J Clin Invest. 2023;133(2).

59. Zhao J, Chen AX, Gartrell RD, Silverman AM, Aparicio L, Chu T, et al. Immune and genomic correlates of response to anti-PD-1 immunotherapy in glioblastoma. Nat Med. 2019;25(3):462–9.

60. Cloughesy TF, Mochizuki AY, Orpilla JR, Hugo W, Lee AH, Davidson TB, et al. Neoadjuvant anti-PD-1 immunotherapy promotes a survival benefit with intratumoral and systemic immune responses in recurrent glioblastoma. Nat Med. 2019;25(3):477–86.

61. Johanns TM, Miller CA, Dorward IG, Tsien C, Chang E, Perry A, et al. Immunogenomics of Hypermutated Glioblastoma: A Patient with Germline POLE Deficiency Treated with Checkpoint Blockade Immunotherapy. Cancer Discov. 2016;6(11):1230–6.

62. Lukas RV, Rodon J, Becker K, Wong ET, Shih K, Touat M, et al. Clinical activity and safety of atezolizumab in patients with recurrent glioblastoma. J Neurooncol. 2018;140(2):317–28.

63. Guo X, Wang S, Wang Y, Ma W. Anti-PD-1 plus anti-VEGF therapy in multiple intracranial metastases of a hypermutated, IDH wild-type glioblastoma. Neuro Oncol. 2021;23(4):699–701.

64. Das A, Sudhaman S, Morgenstern D, Coblentz A, Chung J, Stone SC, et al. Genomic predictors of response to PD-1 inhibition in children with germline DNA replication repair deficiency. Nat Med. 2022;28(1):125–35.

65. Bouffet E, Larouche V, Campbell BB, Merico D, de Borja R, Aronson M, et al. Immune Checkpoint Inhibition for Hypermutant Glioblastoma Multiforme Resulting From Germline Biallelic Mismatch Repair Deficiency. J Clin Oncol. 2016;34(19):2206–11.

66. Suwala AK, Stichel D, Schrimpf D, Kloor M, Wefers AK, Reinhardt A, et al. Primary mismatch repair deficient IDH-mutant astrocytoma (PMMRDIA) is a distinct type with a poor prognosis. Acta Neuropathol. 2021;141(1):85–100.

67. van der Klift HM, Jansen AM, van der Steenstraten N, Bik EC, Tops CM, Devilee P, Wijnen JT. Splicing analysis for exonic and intronic mismatch repair gene variants associated with Lynch syndrome confirms high concordance between minigene assays and patient RNA analyses. Mol Genet Genomic Med. 2015;3(4):327–45.

68. Johannesma PC, van der Klift HM, van Grieken NC, Troost D, Te Riele H, Jacobs MA, et al. Childhood brain tumours due to germline bi-allelic mismatch repair gene mutations. Clin Genet. 2011;80(3):243–55.

