## Supplemental Figures for "Whole genome sequencing-based analysis of genetic predisposition to adult glioblastoma"

**Supplementary Figure 1** Example of command-line arguments that were used to run SAGE in default germline mode

```
java -Xmx24G -jar /opt/tools/sage.jar
-tumor <sampleID>
-tumor_bam /data/input/<sampleID>_dedup.realigned.cram
-hotspots /sage/37/KnownHotspots.germline.37.vcf.gz
-panel_only
-hotspot_min_tumor_qual 50
-panel_min_tumor_qual 75
-hotspot_max_germline_vaf 100
-hotspot_max_germline_rel_raw_base_qual 100
-panel_max_germline_vaf 100
-panel_max_germline_rel_raw_base_qual 100
-ref_sample_count 0
-panel_bed /data/input/ActionableCodingPanel.37.bed.gz
-coverage_bed /data/input/ActionableCodingPanel.37.bed.gz
-high_confidence_bed /giab_high_conf/37/NA12878_GIAB_highconf_IllFB-IllGATKHC-CG-Ion-
Solid_ALLCHROM_v3.2.2_highconf.bed.gz
-ref_genome /reference_genome/37/Homo_sapiens.GRCh37.GATK.illumina.fasta
-ref_genome_version V37
-ensembl_data_dir /ensembl_data_cache/37/
-output_vcf /data/output/<sampleID>.sage.germline.vcf.gz
-threads 16
-write_bqr_data
-panel_only
-hotspot_min_tumor_qual 50
-panel_min_tumor_qual 75
-low_confidence_min_tumor_qual 100
-high_confidence_min_tumor_qual 100
-map_qual_ratio_factor 2.5
-fixed_qual_penalty -15
-min_map_quality 0
-read_edge_factor 0
```

**Supplementary Figure 2** Example of command line arguments that were used to run PAVE in default germline mode

```
java -Xmx24G -jar /opt/tools/pave.jar -sample <sampleID>
-vcf_file /data/output/<sampleID>.sage.germline.vcf.gz
-read_pass_only
-ref_genome /reference_genome/37/Homo_sapiens.GRCh37.GATK.illumina.fasta
-ref_genome_version V37
-driver_gene_panel /gene_panel/37/DriverGenePanel.37.tsv
-ensembl_data_dir /ensembl_data_cache/37/
-mappability_bed /mappability/37/mappability_150.37.bed.gz
-clinvar_vcf /sage/37/clinvar.37.vcf.gz
-blacklist_bed /sage/37/KnownBlacklist.germline.37.bed
-blacklist_vcf /sage/37/KnownBlacklist.germline.37.vcf.gz
-gnomad_pon_filter -1
-gnomad_freq_file /gnomad/37/gnomad_variants_v37.csv.gz
-output_dir /data/output
-output_vcf_file /data/output/<sampleID>.pave.germline.vcf.gz
-threads 8
-log_debug
```

**Supplementary Figure 3** Quality density plot of small variant scores

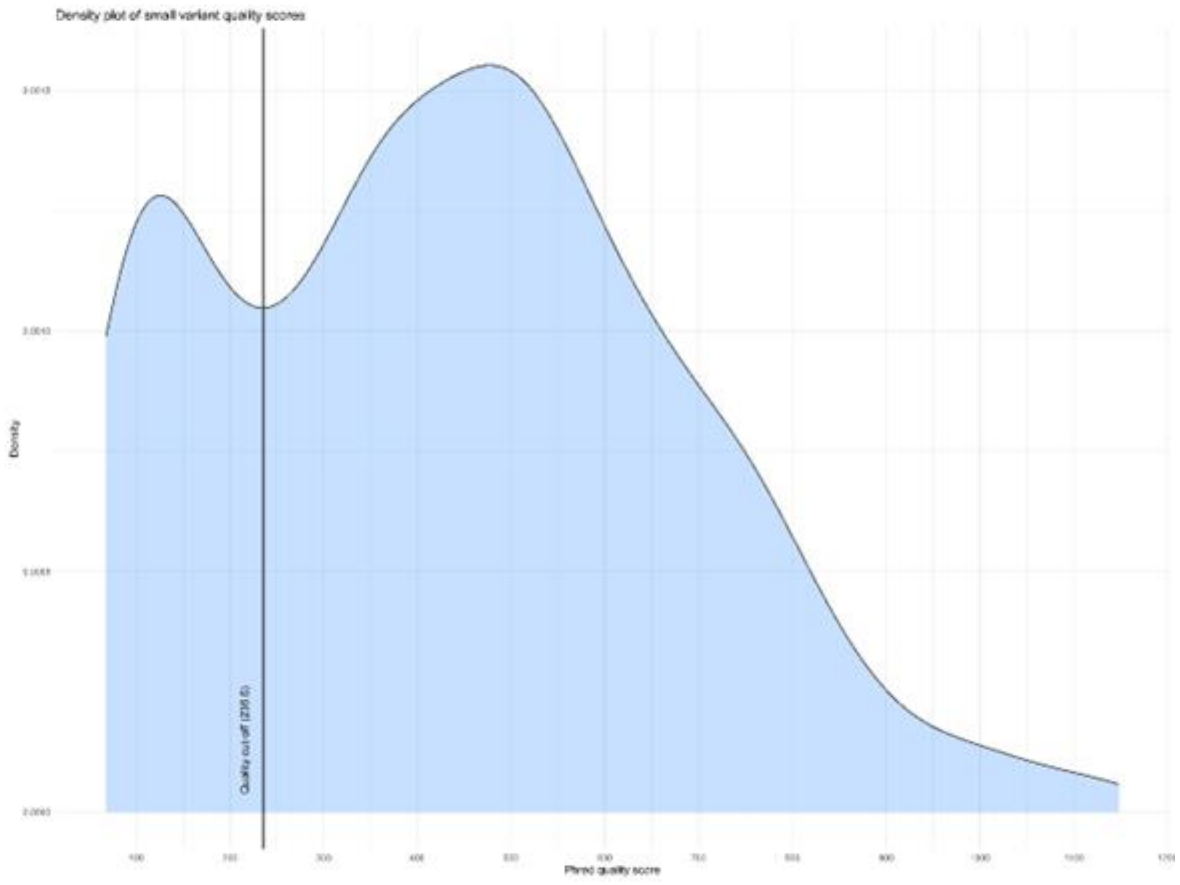

**Supplementary Figure 4** Circos plot (left) and main SBS Cosmic mutational signatures detected (right) of HMF006786A.

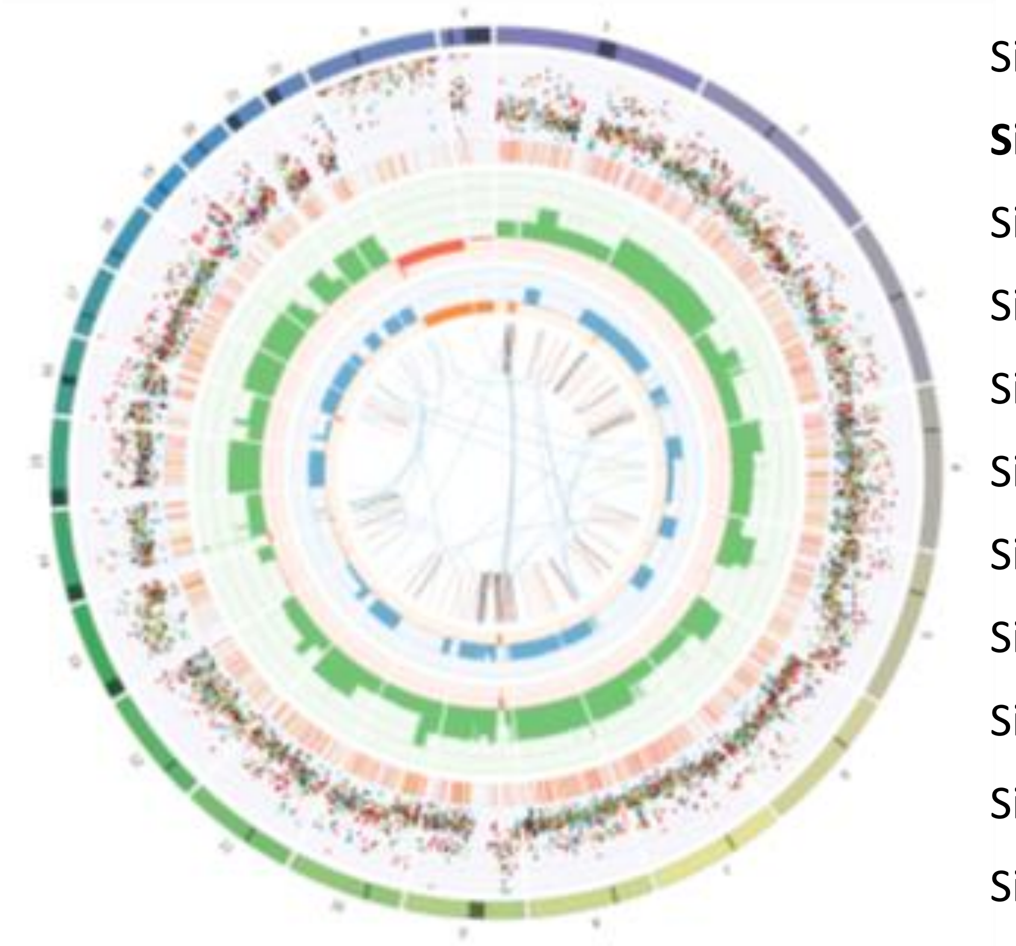

| SIGNATURE | ALLOCATION | PERCENT | |
|---|---|---|---|
| Sig1 | 1431.6 | 19% | |
| **Sig3** | **1974.3** | **26%** | **HRD (Proposed etiology)** |
| Sig9 | 522.7 | 7% | |
| Sig13 | 335.9 | 4% | |
| Sig15 | 803.8 | 11% | |
| Sig16 | 1327.5 | 18% | |
| Sig18 | 180.1 | 2% | |
| Sig26 | 990.1 | 13% | |
| Sig15 | 803.8 | 11% | |
| Sig16 | 1327.5 | 18% | |
| Sig18 | 180.1 | 2% | |
| Sig26 | 990.1 | 13% | |

**Supplementary Figure 5.** Insilico predictions for PMS2:c.825A>G (p.Gly275=) and RNAseq analysis of PMS2 exon 1-12 for HMF000729A (PMS2:c.825A>G & c.325dup)

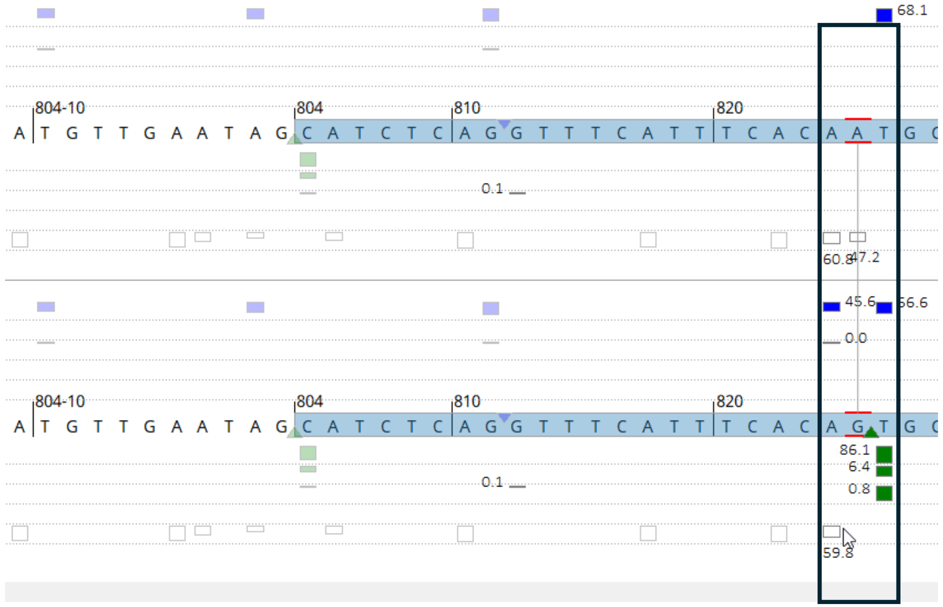


In silico predictions for PMS2:c.825A>G (p.Gly275=) shows an acceptor gain 22 bp upstream of the natural acceptor splice site of exon 8.

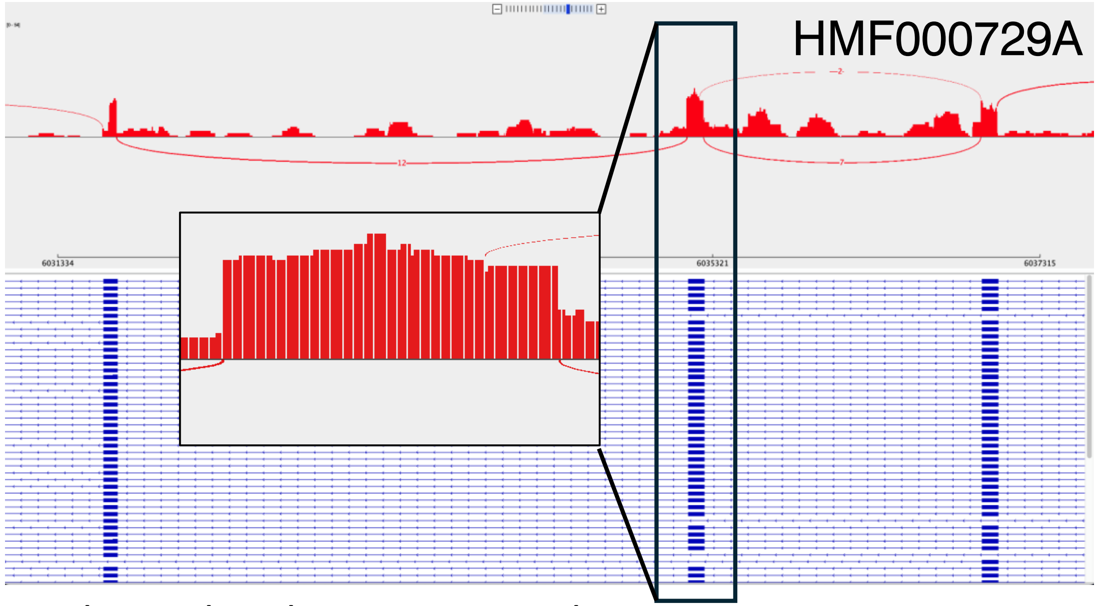


Sashimi plot showing two splice acceptor events at exon 8.
